# Fentanyl Overdose Deaths Decreased Following Scanner Use at Ports-of-Entry

**DOI:** 10.1101/2024.10.16.24315583

**Authors:** Oren Miron

## Abstract

Overdose deaths in the United States surged from 2018 to 2022 due to the smuggling of fentanyl through ports-of-entry. To combat this epidemic, the U.S. used advanced scanners at southern ports-of-entry, which tripled inspections and improved screening, especially since March 2023. Following March 2023, fentanyl domestic seizures decreased by 29% and fentanyl overdose deaths decreased by 35%. Fentanyl overdose deaths decreased in 90% of states and increased only in states that receive fentanyl through ports-of-entry that lack these scanners. If this decrease in overdose deaths continues until November 2025, the death rate would be lower than at the start of the opioid epidemic. These results support urgent funding for advanced scanners at ports-of-entry that lack them.

## Introduction

Fentanyl overdose deaths in the United States (U.S.) surged from 31,335 in 2018 to 73,838 in 2022, due to the smuggling of illicitly manufactured fentanyl through ports-of-entry.^1^ To combat this epidemic, the U.S. installed advanced scanners named Multi-Energy Portals (MEP), which use multiple X-ray energy levels to better identify concealed drugs within cargo vehicles. The MEP also uses lower energy in the driver’s cabin, which allows scanning the truck while it is driven, allowing the scanning of more trucks per hour.^2^ MEP were installed at the ports-of-entry of Nogales Arizona, Santa Teresa New Mexico, and Laredo and Brownsville in Texas. The MEP passed acceptance tests at these ports-of-entry by mid 2023 and increased cargo inspections from 24% to over 80%.^3,4^

The selection of vehicles to inspect with MEP improved significantly with the addition of intelligence on vehicles likely to smuggle drugs since operation Blue Lotus started on March 2023.^5^ This operation brought to the ports-of-entry extensive intelligence personnel and assets from the Department of Homeland Security and led to a surge in fentanyl seizures and arrests. The Blue Lotus operation continued until May 2023, and was followed by operation Blue Lotus 2, and similar operations.^6^ The effect of using MEP on overdose deaths is unknown, largely due the difficulty for medical researchers to access sensitive counter-smuggling information. The information in this study was acquired through months of dialogue with relevant organizations, and extensive searching of archives and press releases.

### Results: Fentanyl seizures at port-of-entry

Data on fentanyl confiscations in the Nogales port-of-entry were manually extracted from 445 press releases made by the port-director, and the date of the MEP initial use was given directly by Customs and Border Protection (CBP).^7^ Fentanyl seizures in Nogales increased in the year after MEP initial use from a yearly average of 2.3 lbs./day to a yearly average of 30.4 lbs./day (13-fold increase; Figure 1).

**Figure 1:**
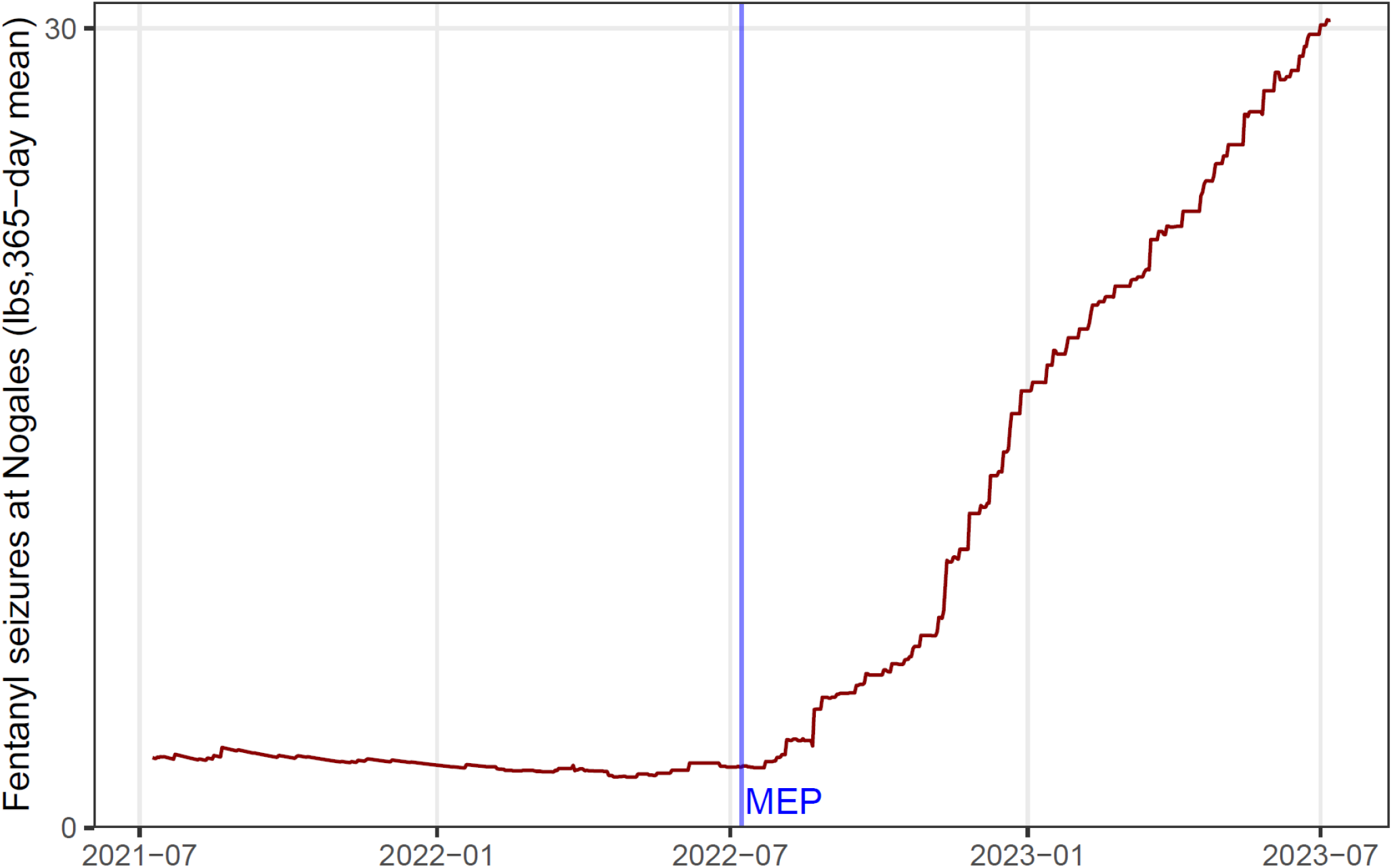
fentanyl seizures at Arizona’s port-of-entry following MEP initial use. Legend: Vertical axis indicates fentanyl pounds seized at Nogales Arizona with a 365-day moving mean. Horizontal axis indicates date, and the blue vertical line indicates the day of Multi-Energy Portal (MEP) initial use.

### Results: fentanyl domestic seizures

The fentanyl ratio among domestic drug seizures was extracted from the National Forensic Laboratory Information System of the Drug Enforcement Administration (DEA) for half year increments from 2013 to 2023. The ratio for the first half of 2024 was given directly by the DEA.^8^ The fentanyl ratio increased consistently in the decade leading up to Blue Lotus, with a 9% increase in the year leading to it, and a 29% decrease in the following year (Figure 2).

**Figure 2:**
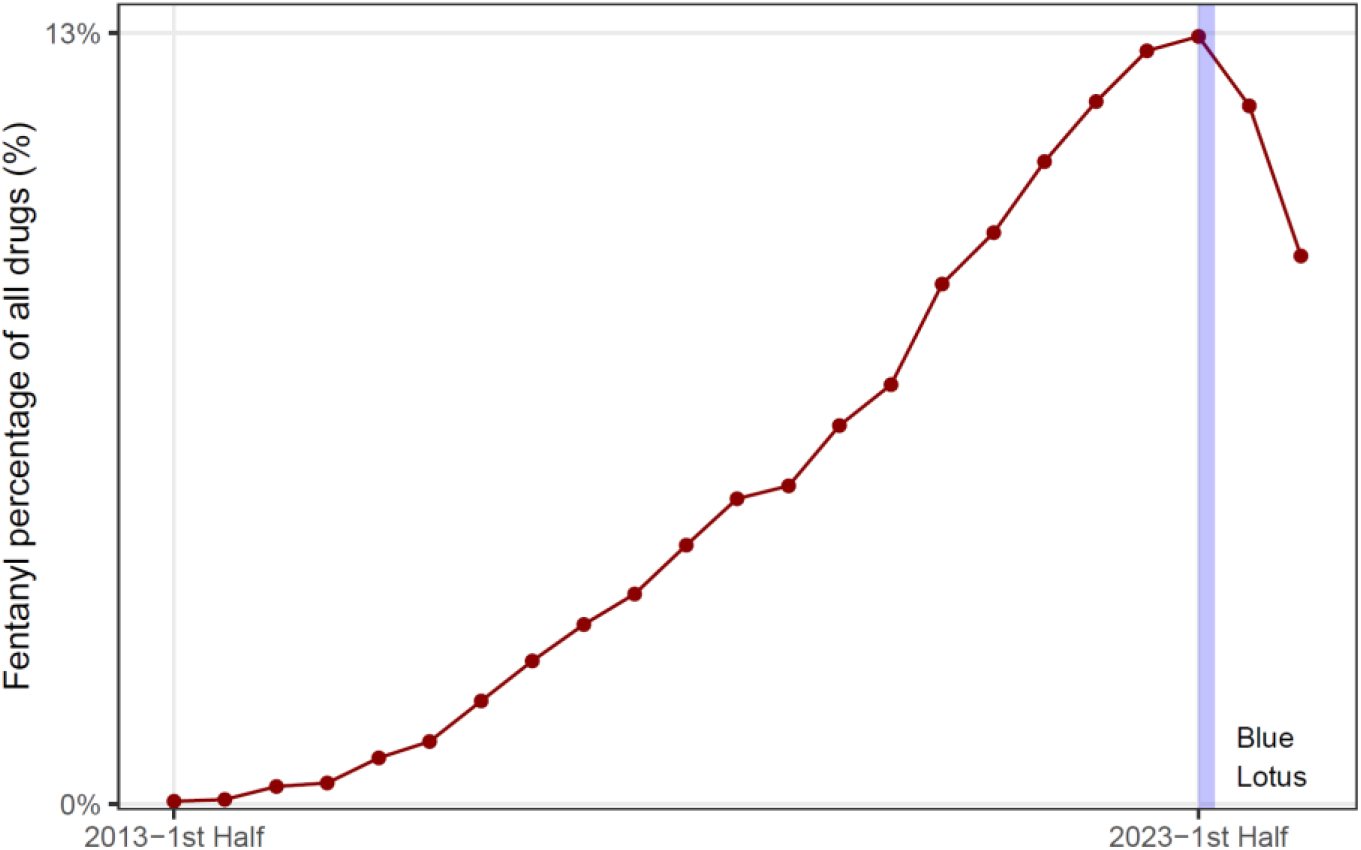
fentanyl ratio among domestic seizures following Blue Lotus. Legend: Y-axis indicates fentanyl ratio among all drugs that the DEA seized domestically and tested through the National Forensic Laboratory Information System. X-axis indicates the half-year of the test. The blue line indicates the start of operation Blue Lotus.

### Results: fentanyl overdose deaths in the U.S

The study extracted monthly fentanyl overdose death counts in the U.S. from the CDC database and adjusted for the number of days per month.^9^ Fentanyl overdose deaths in the U.S. increased from 5,961 in April 2022 to 6,555 in March 2023, when Blue Lotus started (+10%). In the following year, fentanyl overdose deaths decreased to 4,461 in March 2024 (−32%), and in the following month it decreased to 4,260 (−35%). This death count is significantly lower than the predicted deaths for April 2024 (P-value<0.05; -41% decrease; Figure 3).

**Figure 3:**
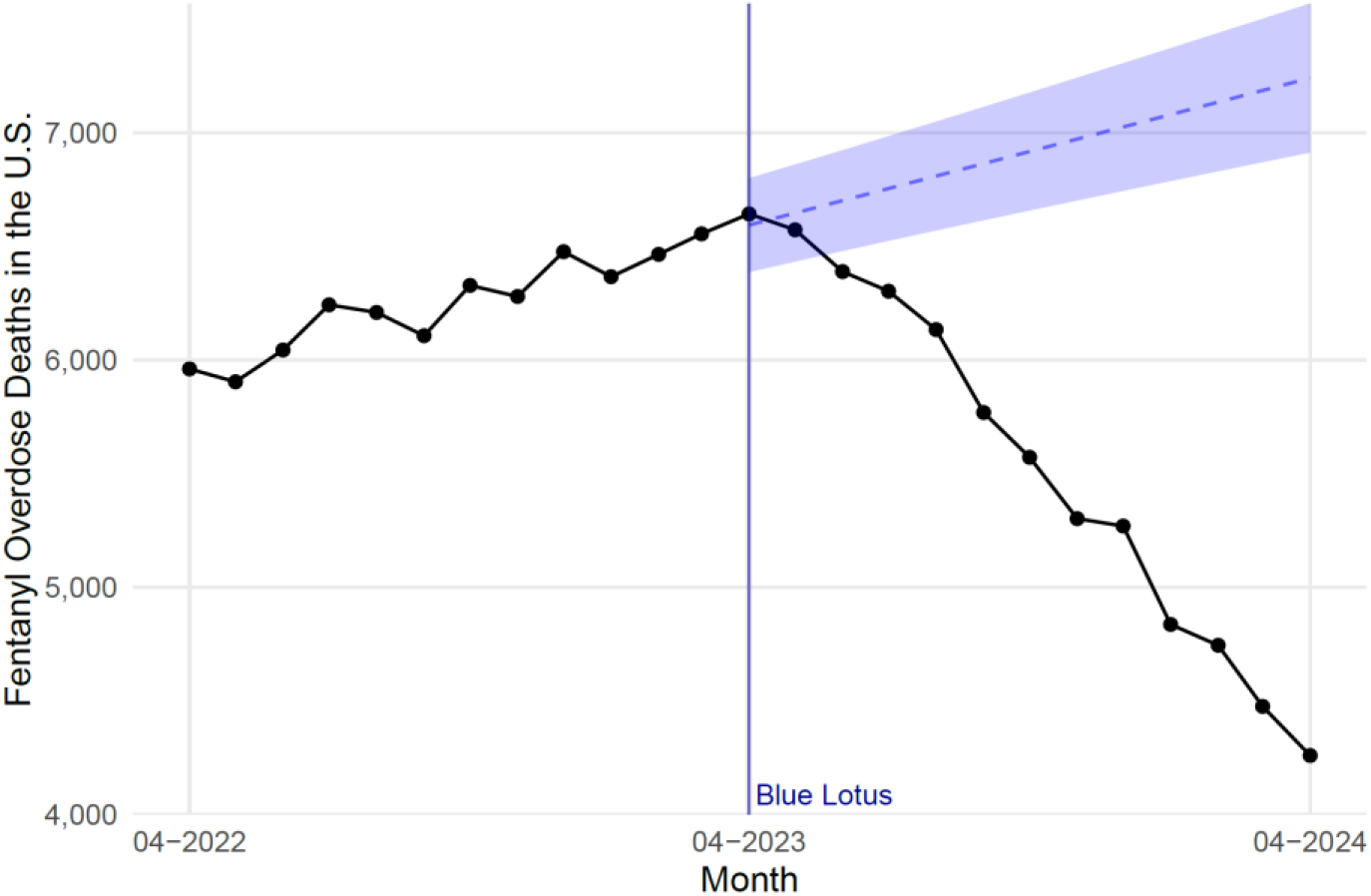
fentanyl overdose deaths in the U.S. following Blue Lotus. Legend: Y-axis indicates monthly fentanyl overdose deaths in the U.S. X-axis represents time. The black line shows observed deaths, the dotted blue line shows predicted deaths based on pre-April 2023 data, and the shaded region represents the prediction’s 95% confidence interval.

### Results: fentanyl overdose deaths by state

Between the first quarters of 2023 and 2024, fentanyl overdose deaths decreased in 46 of 51 states and the District of Columbia, with a median decrease of 27%. The percentage change: - 71 WY, -62 NC, -44 VT, -42 MI, -40 SD, -39 WI, -38 OH, -38 VA, -37 PA, -35 DE, -34 WV, -34 IL, -33 MO, -33 RI, -33 SC, -31 IA, -31 NM, -31 NH, -31 KY, -30 TN, -30 GA, -30 AR, -29 ID, -29 NA, -28 NJ, -27 MA, -27 MN, -27 AZ, -26 AL, -24 OK, -24 NY, -24 MD, -24 LA, -23 IN, -21 CA, - 21 KS, -20 FL, -20 NE, -18 MS, -17 CO, -14 ND, -12 CT, -10 ME, -8 MT, -8 TX, -2 OR, +1 UT, +2 WA, +18 NV, +27 HI, +78 AK. The only 5 states with an increase in fentanyl deaths were in the west and receive fentanyl mainly through California ports-of-entry that lack MEP (Figure 4).

**Figure 4:**
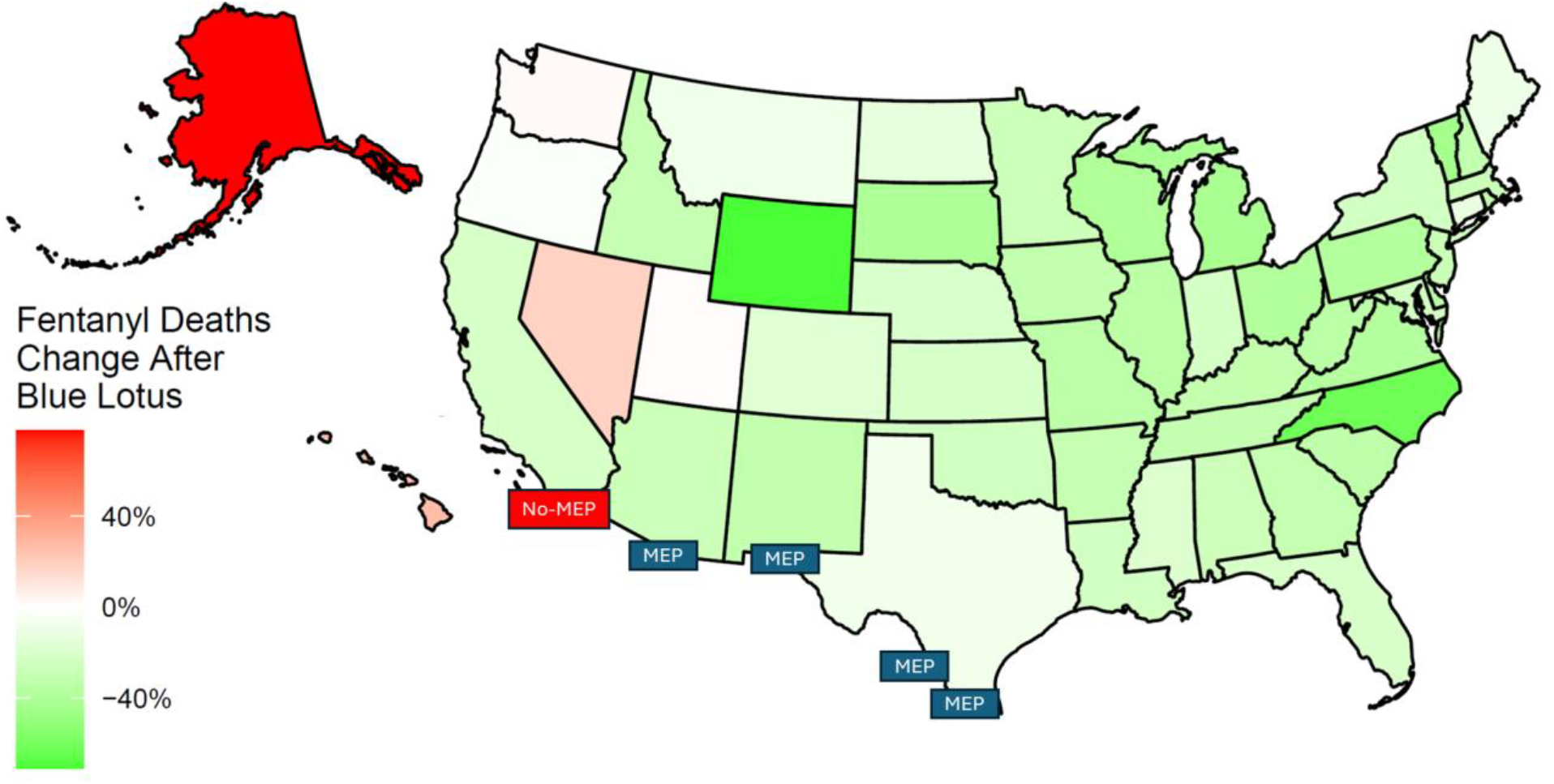
change in fentanyl overdose Deaths by State following Blue Lotus. Legend: The map shows the percentage change in fentanyl overdose deaths by state between the first quarter of 2023 and the first quarter of 2024. Around the end of the first quarter of 2023, the U.S. used Multi-Energy Portal (MEP) at ports-of-entry in Arizona, New Mexico, and Texas, but not in California. The color gradient indicates the change, with red representing increases and green representing decreases. Ports of entry with MEP are indicated in a blue rectangle and the ports-of-entry of California without MEP are in red.

### Results: opioid overdose deaths

The opioid overdose death rates per month were analyzed from 1999 to 2024. The lowest rate was 2/million population in October 2000, which increased 11-fold until Blue Lotus in March 2023. In the year following Blue Lotus, opioid overdose deaths decreased by 31%, and the following month the decrease was 34%. The linear model based on this decrease predicts that the rate would reach 2/million in November 2025 (Figure 5).

**Figure 5:**
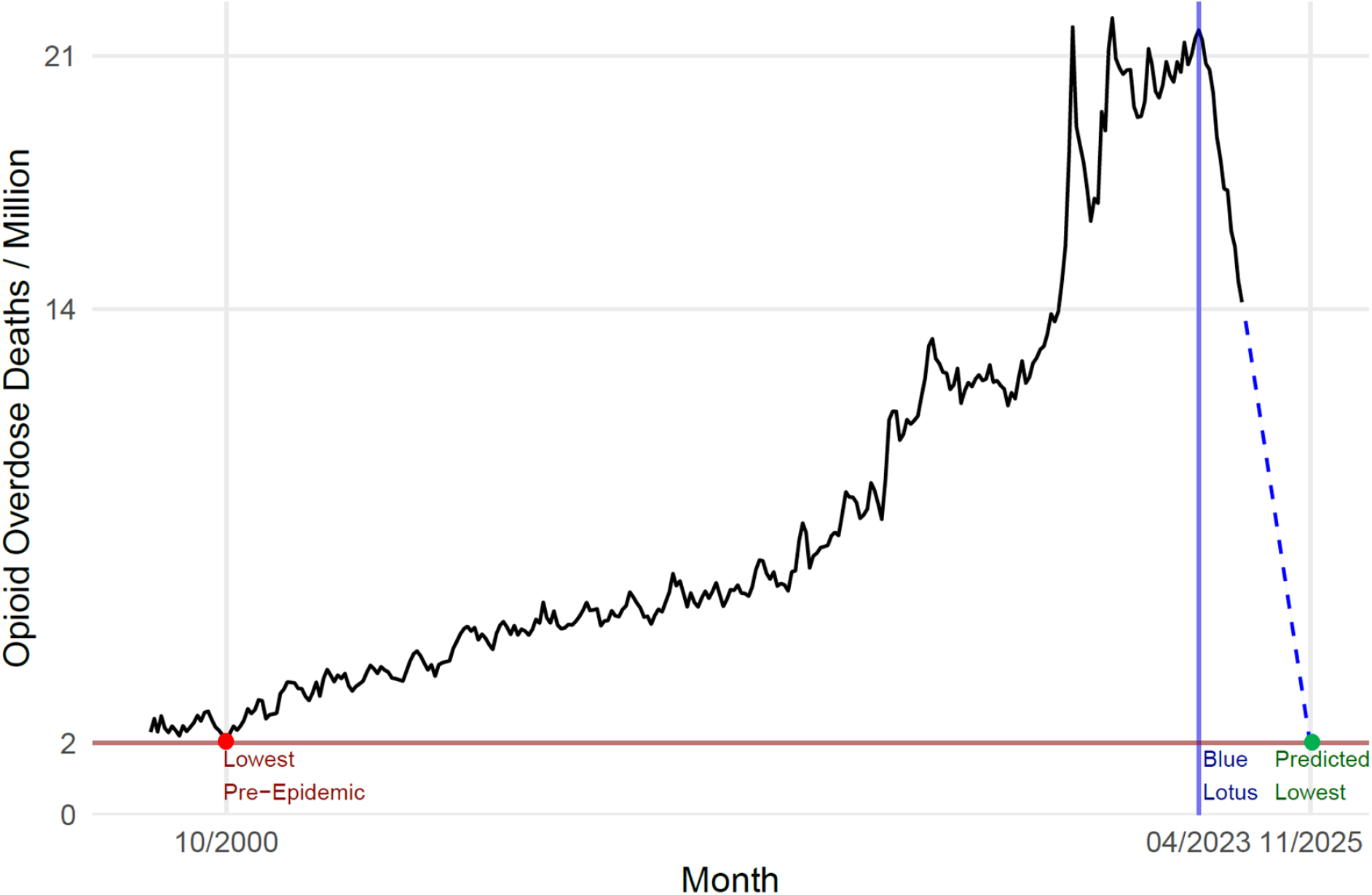
opioid overdose deaths in the U.S., 1999-2024, with prediction to 2025. Legend: Y-axis indicates opioid overdose deaths per 1,000,000 United States population. X-axis indicates the month of death. The red line indicates the lowest rate before the opioid epidemic started to increase. The blue line indicates Operation Blue Lotus, and the dotted line indicates the predicted rate based on the linear model of the rate after Blue Lotus.

## Discussion

These findings indicate that MEP use was followed by a sharp increase in fentanyl seizures at ports-of-entry, and sharp decrease in fentanyl availability and overdose deaths. A similar trend was found in opioid overdose deaths, with the decreasing trend predicted to reach pre-epidemic rates by Nov. 2025. These monthly decreases in overdose deaths are three times larger than the decrease in annual deaths that was widely reported, which highlights the importance of tracking fentanyl deaths monthly as well as annually.^10^

While fentanyl overdose deaths fell in most states, they increased only in states that receive fentanyl through California’s ports-of-entry, which did not receive MEP due to delay in funding. This pattern might explain reports of decreased fentanyl availability across the U.S. except around the west coast.^11^ These findings suggest a need to urgently fund MEP in ports-of-entry in California and across the U.S.

## Data Availability

All data produced are available online at: https://wonder.cdc.gov/controller/saved/D176/D408F378

## Acknowledgements

I would like to thank the law enforcement experts, who provided valuable insights that made this study possible. I would also like to thank those on the front lines and across America that are fighting the opioid epidemic and saving countless lives.

